# The Pathology, Alzheimer’s and Related Dementias Study (PARDoS): Design and Characteristics of the First 4700+ Brazilian Participants

**DOI:** 10.1101/2025.04.12.25325490

**Authors:** Jose M. Farfel, Ana W. Capuano, Sukriti Nag, Maria Carolina M. Sampaio, John Gibbons, Robert S. Wilson, David A. Bennett

**Affiliations:** Rush Alzheimer’s Disease Center, Rush University Medical Center, Chicago, IL, USA; Department of Pathology, Rush University Medical Center, Chicago, IL, USA; Instituto de Assistência Médica ao Servidor Público Estadual, São Paulo, SP, Brazil; Department of Neurological Sciences, Rush University Medical Center, Chicago, IL, USA; Department of Behavioral Sciences, Rush Medical College, Chicago, IL, USA

## Abstract

The Pathology, Alzheimer’s and Related Dementias Study (PARDoS) is a community-based clinical-pathologic study of aging and dementia in a large and diverse sample of Brazilians. Its long-term objective is to identify the environmental, genetic and molecular drivers of common conditions across the adult life span with an emphasis on Alzheimer’s Disease and Related Disorders clinical and neuropathologic traits. From July 31^st^ 2021 through February 11^th^, 2025, more than 4,700 brains were collected at two autopsy centers and a major hospital system in the State of Sao Paulo, Brazil. Samples of other organs are also being collected. Their mean age was 71.7 years (range 18-106), 40.2% were Black/Mixed, 52.7% were male, their mean education was 6.3 years (range 0-25). Among those aged 65+, 32.9% had dementia and 18.8% had mild cognitive impairment. Neuropathologic data collection is ongoing. PARDoS fills several major gaps among clinical-pathologic studies given the large numbers and its unique age and education range, and socioeconomic status, race, sex, and other organ collection. Here we present the study design, demographic characteristics of the first 4,790 autopsied participants, and clinical characteristics of the first 4,283 with informant interviews.

## Introduction

Alzheimer’s Disease and Related Disorders (ADRD) are major public health priorities in the developed and developing world.[1–12] Clinical-pathologic studies of aging and dementia remain an important part of the research landscape as many brain pathologies cannot be interrogated in living persons. Further, the molecular events leading to ADRD pathologies and linking them to subsequent dementia can only be studied in detail in brain tissue at this time. Most clinical-pathologic studies are of patients seen in tertiary care dementia clinics.[13,14] While this is necessary for rare dementias [15], it has limitations for common diseases such as Alzheimer’s dementia in which tertiary care attracts atypical cases as we and others have shown.[16–18] Yet, there are relatively few community-based clinical-pathologic studies of aging and dementia [19–29], including those led by our group at the Rush Alzheimer’s Disease Center (RADC) [30–32], which are composed mostly of very-old, highly educated, non-Latinx white women. An exception is a another clinical-pathologic study in Brazil which collected about 1900 brains from older Black and White decedents over the past two decades.[33,34] Given the complexity of ADRD, there is a clear need for studying brains from persons with more diverse age, sex, race, education and socioeconomic profiles. Further, given the increasing interest in the exposome and other interactions between the brain and the body [35–39], the ability to obtain other internal organs for study paired with the brain is also an ongoing need. Brazil is the ideal location to perform a study to fill these gaps.

PARDoS is based at the Instituto de Assistência Médica ao Servidor Público Estadual (IAMSPE), a public tertiary-care hospital located in the city of Sao Paulo which provides healthcare to the public servants of the State of Sao Paulo and their families and started brain collection on July 31, 2021. The study collects brains at two autopsy services in the State of Sao Paulo as well as the Hospital Servidor Público Estadual (HSPE) of IAMSPE. Here we report the design and baseline demographic characteristics of the first 4,790 PARDoS autopsies and the clinical characteristics of the first 4283 participants collected through February 11, 2025.

## Methods

### Study participants

Decedents aged 18 years or older with a legal representative are eligible for PARDoS. Recruitment is designed to enrich for African Ancestry by prioritizing, proxy-report Black/Mixed and Whites decedents with 8 or fewer years of education. Decedents born in one of the three Southern states of Brazil of Paraná, Santa Catarina and Rio Grande do Sul are low priority as the population in those states have limited African ancestry.

PARDoS was approved by an Institutional Review Board of Rush University Medical Center, and as a new project by the IAMSPE local ethics committee (CEP-IAMSPE) on May 12, 2020 and by the Comissão Nacional de Ética em Pesquisa (CONEP), the Brazilian national ethics committee on September 17, 2020. Due to the COVID pandemic, the first brain was not obtained until mid-2021. PARDoS collects data and biospecimens at three sites (Figure 1).

**Figure 1.**
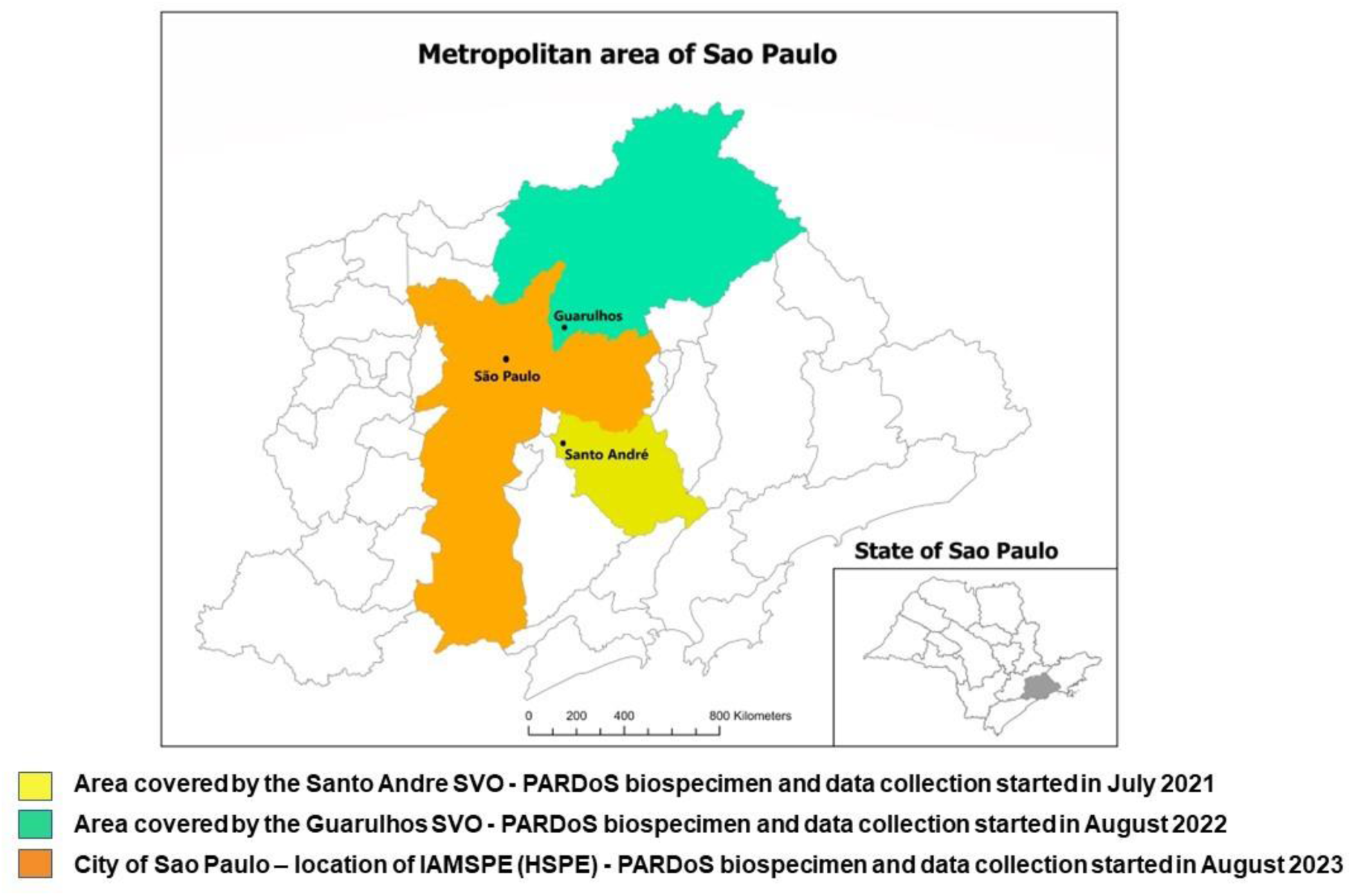
PARDoS workflow at the recruiting sites.

A law passed in Brazil in 1939, requires a death certificate to include causes of death signed by a physician. Since many Brazilians do not have a physician to sign the death certificate, these decedents are taken to a designated autopsy service in their catchment area where a full-body autopsy is done to determine the causes of death. These autopsy services are for decedents who die from natural causes and are distinct from the forensic autopsy services.

The Santo Andre Autopsy Service (Santo Andre SVO), southeast of the city of Sao Paulo, covers a population of more than 1 million people. PARDoS started July 31, 2021 at the Santo Andre SVO. The Guarulhos Autopsy Service (Guarulhos SVO), northeast of the city of Sao Paulo, covers a population of about 1.5 million people. PARDoS started August, 2022 at the Guarulhos SVO. A new autopsy suite was built for PARDoS in HSPE and PARDoS started there on August, 2023. At HSPE, all patients have a physician to sign a death certificate, these autopsies are for research only.

After presenting background information about aging and AD, PARDoS nurses ask a legal and knowledgeable representative from eligible decedents to sign an Informed Consent detailing that a data repository will be kept at the Rush Alzheimer’s Disease Center (RADC) and a biorepository at IAMSPE.

Representatives unable to understand the terms of the consent or who are overly distraught are excluded. Cases are ineligible if the brain shows advanced signs of autolysis, or the autopsy service pathologist elects to retain the brain for further investigation, or if the body is later referred to the Forensics.

From July 31, 2021 to February 11, 2025, 10,123 representatives of eligible decedents for PARDoS were approached, of whom 8.3% were not eligible for consent. Consent was obtained from 5307 (57.2%) of the eligible representatives. Of those with consent, 517 (9.7%) decedents were ineligible because the brain could not be obtained for research. In less than 3.5 years, PARDoS enrolled 4,790 decedents. Figure 2 illustrates the study flow for 65+ and 18-<65 participants, including2563 from the Santo Andre SVO, 1755 from the Guarulhos SVO, and 472 from HSPE. Figure 3 shows the rate of enrollment per site from July 2021 to February 2025. The mean age of death for these cases was 71.7 years (SD: 15.1 years, range 18 to 106), 32.7% were age 18-<65 years, 52.7% were male, 40.2% were Black/Mixed, 58.1% were Whites and 1.7% other races, primarily Asian (i.e., Japanese) and the mean education was 6.3 years (SD: 4.3 years). Figure 4 illustrates the race distribution at the three enrollment sites.

**Figure 2.**
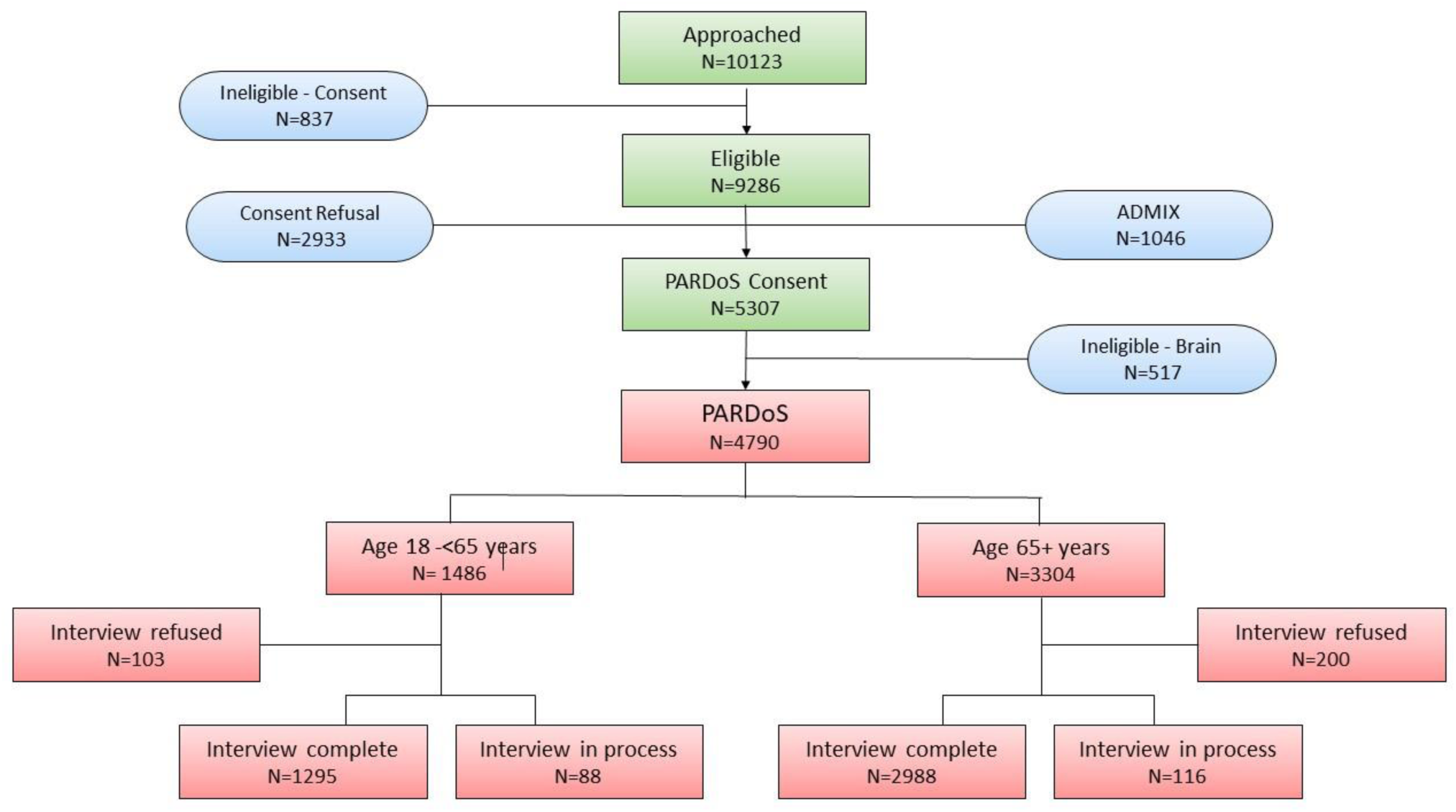
PARDoS recruitment and enrollment flowchart.

**Figure 3.**
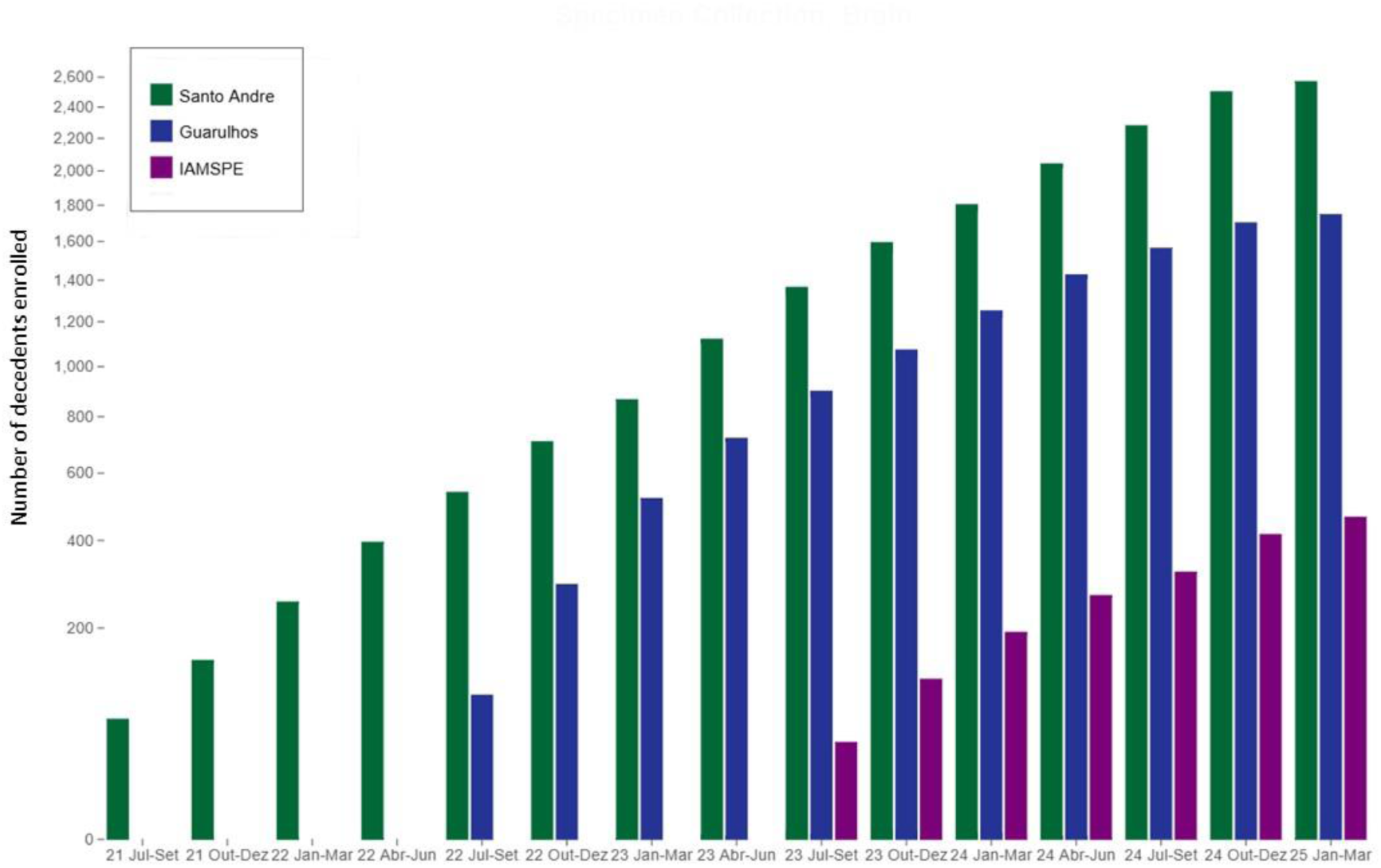
Rate of PARDoS enrollment in the three sites from January 2021 to February 2025.

**Figure 4.**
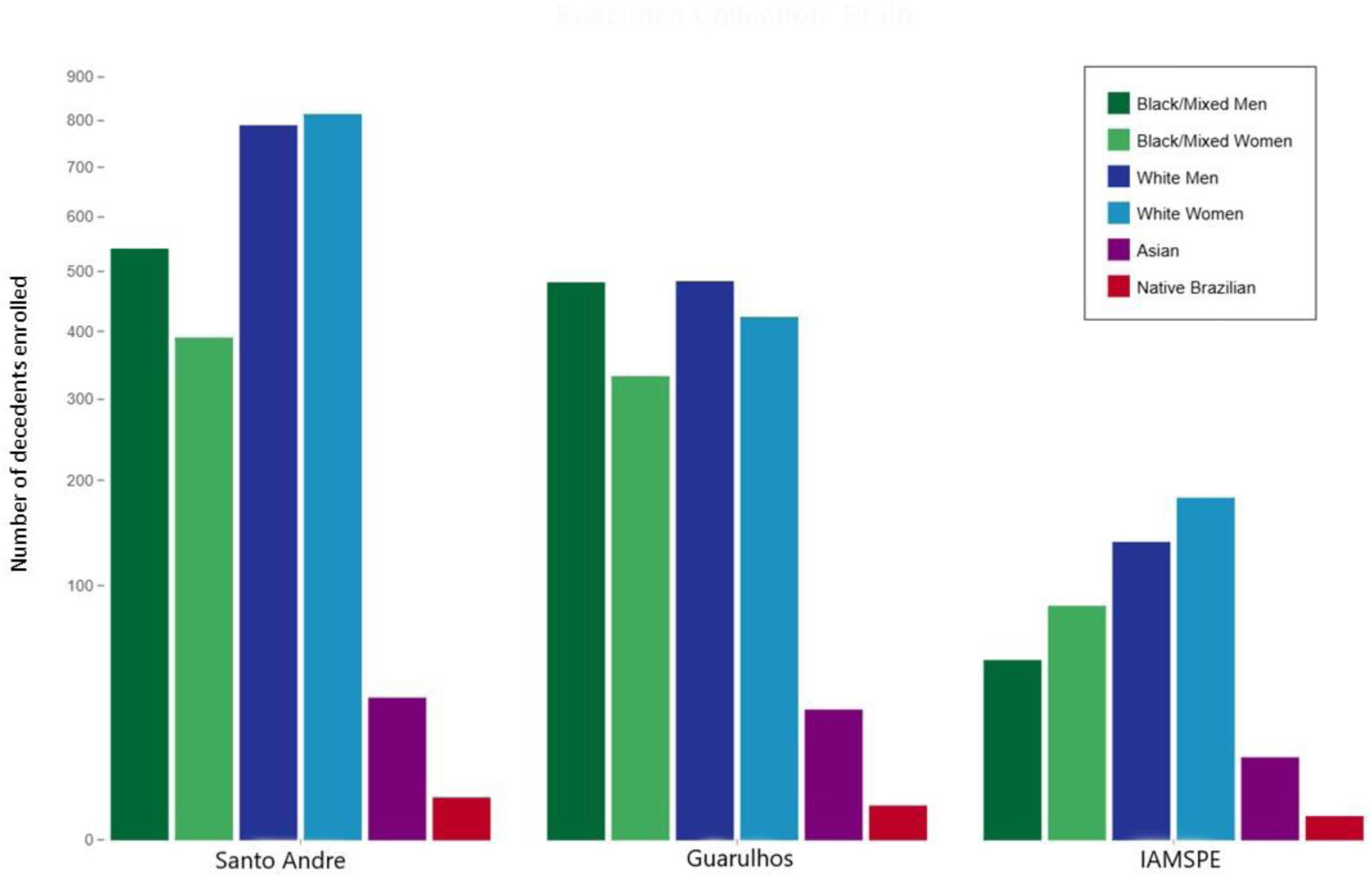
Race distribution of PARDoS participants from July, 2021 to February, 2025.

### Data Management

Briefly, a secure website hosted at Rush University Medical Center (RUMC) provides data entry and workflow processing for PARDoS. The server is located in the Rush Data Center protected by the RUMC enterprise firewall. All PARDoS data are stored in a relational database behind the RUMC firewall. User accounts for PARDoS website are created for study staff with access controlled by role on project. Data entered in Portuguese appear in English in the RADC database in real-time. The only data stored at HSPE are the study consents The data workflow is outlined in Figure 5. More information is in supplementary material.

**Figure 5.**
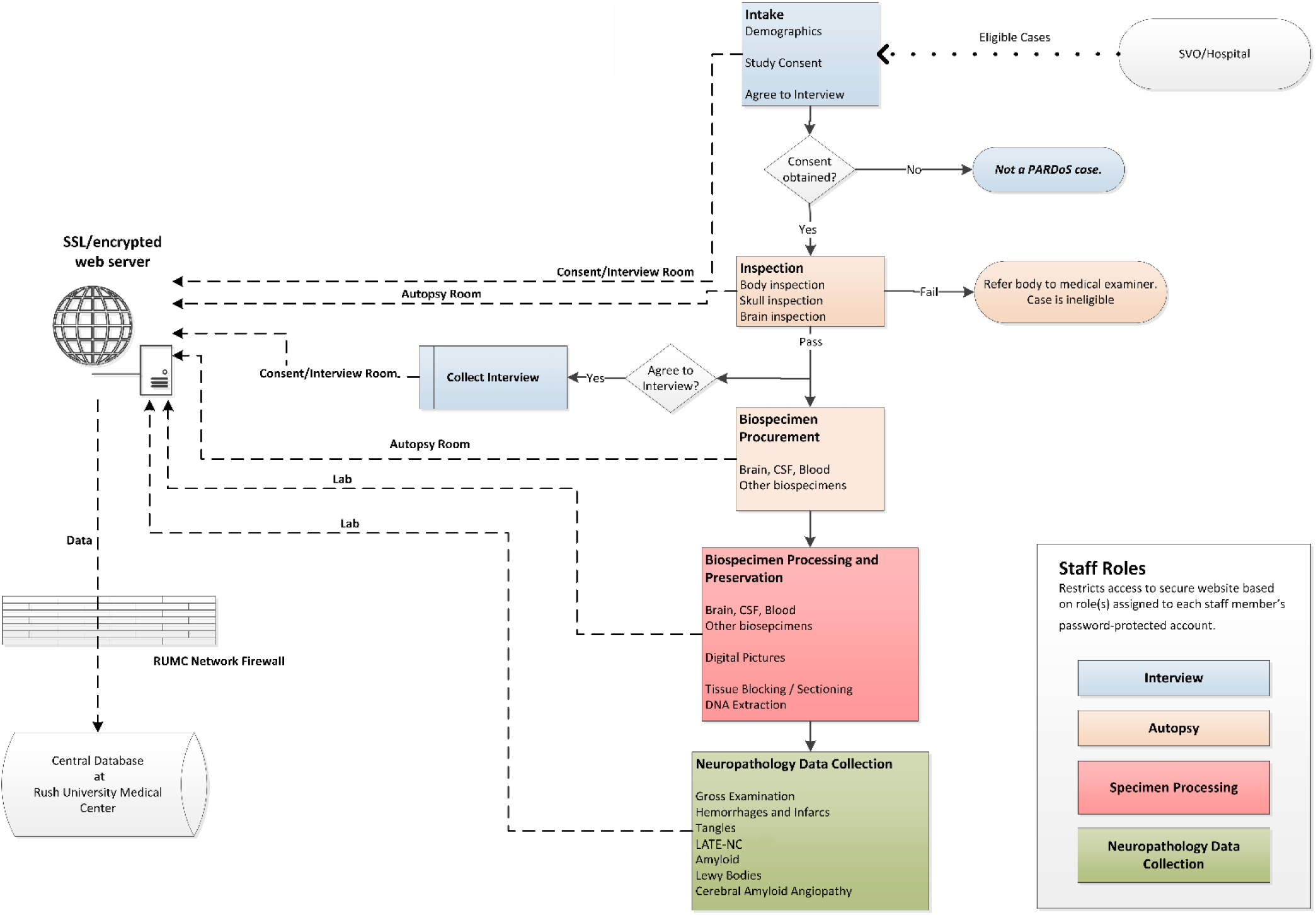
PARDoS data management flowchart.

### Informant interview

Nurse interviewers conduct an interview with one or more informants to obtain ante-mortem information about the decedent. The interview is often conducted on-site immediately following consent; or may be conducted by phone in the days or weeks following the autopsy at a time the informant deems more convenient. Therefore, interview numbers lag behind autopsy numbers. Select components of the clinical interview can be found in Table 1.

**Table 1.**
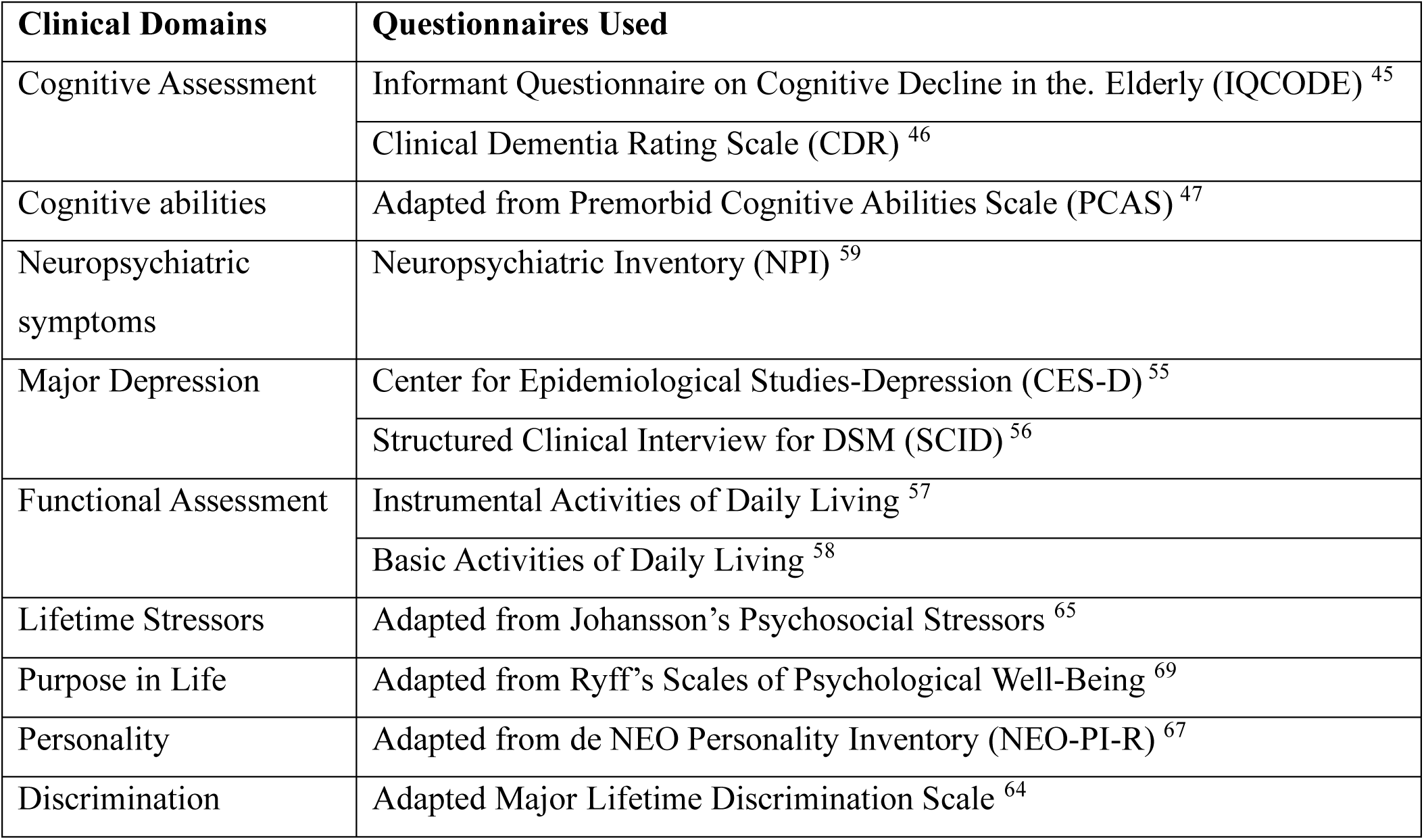
Summary of questionnaires included in the PARDoS clinical interview.

### Subject identification

The complete name of the decedent, date of birth, sex, race classified as Black, Mixed, White, Asian, and Native-American is obtained.[40] Here, we followed multiple Brazilian studies and guidelines that combine Blacks and the Mixed.[41,42]. Education was reported as the number of years of formal education up to 25 years. Ethnicity is not a construct used in Brazil. However, Brazilians in the USA are classified as non-Hispanic Latino.

### Informant identification

We collect the age, sex, and education of the primary informant. Their relationship with the decedent and level of contact with the decedent are classified as daily, weekly, monthly or annual contact. The number of years the informant knew the decedent is also recorded.

### Socioeconomic status

Socioeconomic status (SES) is determined using the Brazilian Association of Research Companies scale which characterizes the number of different household goods and the level of education of the principal householder.[43] The scale uses strata A through E with A and B being the most privileged and D and E the most vulnerable. The C strata is the most common group. We also use the MacArthur Scale of Subjective Social Status to measure proxy subjective impression of the social status of the decedent using a stepladder image.[44]

### Decedent Medical History

The presence of common chronic medical conditions, including memory impairment, dementia, stroke, Parkinson’s disease, cancer, vascular disease (e.g., claudication, heart attack), vascular risk factors (e.g., arterial hypertension, diabetes) are recorded. We also enquire about smoking habits and history of alcohol and drug abuse.

### IQCODE and Clinical Dementia Rating Scale

The 26-item Informant Questionnaire of Cognitive Decline in the Elderly (IQCODE) is a measure of cognitive decline that compares the cognitive status proximate to death with the status the participant had 10 years earlier.[45] The IQCODE final score is calculated as the sum of all the item scores divided by the total number of applicable items to adjust for missing data.

The Clinical Dementia Rating Scale (CDR) is used to determine the presence and severity of cognitive impairment proximate to death.[46] The CDR structured questionnaire for the informant is used for each of the six domains (memory, orientation, judgment and problem solving, community affairs, home and hobbies, personal care). Each domain is scored by an algorithm based on responses on a 5-point scale: 0, no impairment; 0.5, questionable impairment; 1, mild impairment; 2, moderate impairment; and 3, severe impairment (personal care is scored on a 4-point scale combining 0 and 0.5 ratings). The domain scores are converted by an algorithm into an overall rating, which is reviewed by a clinician who either agrees or changes the final score

### Cognitive abilities and Literacy

Lifetime acquired cognitive skills are assessed using the Premorbid Cognitive Skills Questionnaire (PCSQ), a 26-item questionnaire adapted from the Premorbid Cognitive Abilities Scale (PCAS) and items from other extant instruments.[47–54] The final PCSQ score is obtained by the sum of all positive answers to the items, varying from 0 to 26 points.

Three forms of literacy are ascertained. Verbal literacy includes 10 questions, i.e., reading and understanding short phrases. Numeracy includes 3 questions: multiplication, percentages, and use of a calculator. Music literacy is assessed by a single question regarding music reading.

### Depressive Symptoms and Major Depression

Mood and depressive symptoms are examined with the 8-item questionnaire from the Center for Epidemiologic Studies Depression Scale (CES-D) [55] and major depression episode in the past is examined with the Structured Clinical Interview for DSM (SCID) questionnaire.[56] Both questionnaires were adapted for use with an informant.

### Diagnostic Classification

Clinical diagnoses are designed to capture both common and rare conditions that may result in a natural cause of death in adults aged 18+. Thus, in addition to the dementia and mild cognitive impairment (MCI) diagnoses that are common in older persons, we also diagnose cognitive impairment with no dementia (CIND) to capture persons with lifelong cognitive impairment. Dementia and MCI are further classified as Alzheimer’s dementia and amnestic MCI based on the presence of memory impairment. Non-Alzheimer’s dementia or amnestic MCI is further diagnosed as vascular cognitive impairment, Parkinson’s/Parkinsonism related cognitive impairment and other. No cognitive impairment (NCI) is present in the absence of the other three classifications. Diagnoses are rendered using a 2-stage process. First an algorithm combines the IQCODE as a measure of cognitive decline and the CDR as a measure of cognitive impairment to provide a putative diagnosis of dementia, Alzheimer’s dementia, MCI and amnestic MCI. Then an experienced clinician reviews selected relevant information available in the informant interview and confirms or modifies the algorithm results rendering a final clinical diagnosis.

Other potential contributors to cognitive impairment are recorded including, seizures, brain tumors, schizophrenia, bipolar disease, drug and alcohol abuse, mental retardation, autism, microcephaly, Down Syndrome, anoxic encephalopathy, multiple sclerosis, dialytic chronic kidney disease and HIV infection.

### Functional assessment

Instrumental activities of daily-living (IADL) are assessed with the Lawton IADL scale, an 8-item questionnaire.[57] Basic activities of daily-living (BADL) are assessed using the Katz Index of Independence, a 6-item questionnaire.[58]

### Neuropsychiatric symptoms

The Neuropsychiatric Inventory assesses behavioral and psychological symptoms.[59] Neuropsychiatric symptoms are grouped into four clusters: agitation, affect/apathy, psychosis, and behavioral problems as previously described.[60]

### Psychosocial assessment

The interview includes the Three-Item Loneliness Scale obtained from the Revised UCLA Loneliness Scale.[61] The final score is the sum of all items. Higher scores indicate greater degrees of loneliness. We also ask 6 questions regarding social support which quantify the number of children, family members, and friends the decedent had contact with over the last month.

A 7-item questionnaire is used to determine if the decedent was a victim of discrimination or unfair treatment over the life course.[62] We adapted 3 questions from the Everyday Discrimination Scale intended to capture chronic experiences of unfair treatment in day-to-day situations, and 4 questions from the 11-item Major Lifetime Discrimination scale to document discrimination in specific instances.[63,64] Adult exposure to stressors is examined with the 10-item questionnaire developed by Johansson et al.[65] The 10-item Adverse Childhood Experience (ACE) Questionnaire is used for documenting childhood stressful experiences.[66]

Conscientiousness and neuroticism personality traits are determined with 6 questions for each trait based on the Revised NEO Personality Inventory (NEO-PI-R).[67] Purpose in life is documented by 6 questions extracted from published references.[68,69]

### Neuropathologic examination

#### Post-Mortem Procedures

Post-mortem tissue procurement is performed by trained pathology technicians and largely conforms to procedures used at the RADC for the Religious Orders Study and Rush Memory and Aging Project (ROSMAP) with modifications to accommodate for the scale of this study which currently yields about 6 autopsies per day.[32] Procedures include those recommended by the National Alzheimer’s Disease Coordinating Centers (NACC) and recent updated guidelines for the pathologic diagnosis of AD and related disorders.[70–72]

Technicians measure head circumference, and take 20ml of arterial blood, stored in EDTA and serum tubes. The skull is opened and dura detached. Digital photographs of whole brain and abnormal findings are taken. 10-20 mL of CSF is extracted from the occipital horn. After brain removal, the olfactory bulbs and tracts are removed. One olfactory bulb/tract is frozen and stored at −80°C and the other is fixed in 10% buffered formalin. The brainstem and cerebellum are separated from the cerebral hemispheres which are then separated by a sagittal midline cut. The hemisphere with less or no gross pathology is cut into 1 cm slabs using a plexiglass jig and the slabs are snap frozen on metal plates over dry-ice and stored at −80°C. The other hemisphere is fixed in 10% buffered formalin A midline cut separates the two cerebellar hemispheres which are further cut into 1 cm slabs. Slabs from one cerebellar hemisphere are fixed in formalin while slabs from the other hemisphere are frozen and stored at −80°C. A medial sagittal cut is made to divide the brainstem in two. One half of the brainstem is frozen and the other is fixed. Digital photographs are taken of all cerebral and cerebellar slabs. The size, age, and location of all macroscopic infarctions and other pathology are photographed and recorded. The post-mortem interval, which is the time in hours and minutes between the time of death and the time of brain fixation and freezing, is recorded.

After about 3-4 weeks, trained research assistants cut the fixed hemisphere into 1 cm slabs using a plexiglass jig, take digital pictures of the slabs and dissect the tissue to obtain brain blocks from at least 15 anatomic regions of interest: Midfrontal cortex, .Middle temporal gyrus, Anterior temporal pole, Inferior parietal cortex, Occipital cortex, Amygdala, Entorhinal cortex, Anterior and Mid-hippocampus, Basal ganglia, Anterior thalamus, Cerebellar hemisphere, Midbrain, Anterior watershed, and Olfactory bulb/tract. Samples of macroscopic infarctions and other noted pathology are taken.

#### Neuropathologic data collection procedures

All sections are stained with Hematoxylin & Eosin and selected slides for IHC. Specific phosphorylated, monoclonal antibodies identical to those used in ROSMAP, directed against amyloid-β, PHFtau, pTDP-43 and α-synuclein are used.

Post-mortem data collection is performed by trained research associates under the supervision of two study neuropathologists. Major pathologies such as AD neuropathologic change, cerebral amyloid angiopathy (CAA), vascular disease, Lewy Body Disease (LBD), hippocampal sclerosis (HS), and limbic predominant age-related TDP-43 encephalopathy-neuropathologic change are assessed.[73–81] Neuropathological data will be published separately.

### Other organs

The autopsy performed to determine the cause(s) of death in the autopsy services is a full-body examination. PARDoS is approved to collect and examine ADRD pathology in a variety of bodily organs. Other analyses are determined by the needs of separately funded studies. For example, we collected teeth and skull bone samples for early-life and lifetime metal exposure. We conducted a pilot study with lung and bronchus for carbon. We are collecting pericardial and abdominal fat and liver and planning for a pilot study to examine persistent organic pollutants. There are plans to collect other tissues for microplastics.

### Molecular Genomics

PARDoS is currently funded to generate *APOE* and genome-wide data from brain genomic DNA to identify genomic variants of African ancestry associated ADRD neuropathologic and clinical traits. PARDoS is also approved for RNA sequencing, high-throughput proteomics, and DNA methylation techniques We currently have pilot funding for bulk RNAseq, whole genome DNA methylation, and metabolomics in select participants.

### Aging and Dementia Multiracial Genetic Cohort (ADMIX)

To augment power for genomic analyses of clinical traits, we initially asked families of those who refused brain collection to participate in the clinical interview and collection of a blood sample and named this study ADMIX. No data from ADMIX are presented in this manuscript (see supplementary text).

### Study of Ancestry Neurodegenerative Diseases and Stroke (SANDS)

Prior to the launch of PARDoS, we conducted SANDS from November 2016 to July 2018 at a different institution in Sao Paulo. No data from SANDS are presented in this manuscript (see supplementary text).

### Data Analysis

No formal statistical tests of the data from the two groups were made, as the purpose of this manuscript is to present the study design, demographic and clinical characteristics of the sample.

## Results

We present the demographic and clinical characteristics of the sample by White and Black/Mixed decedents. To compare the sample characteristics with other clinicopathological studies of older adults we present the results separately for decedents ages 65+ and <65 in Table 2).

**Table 2.**
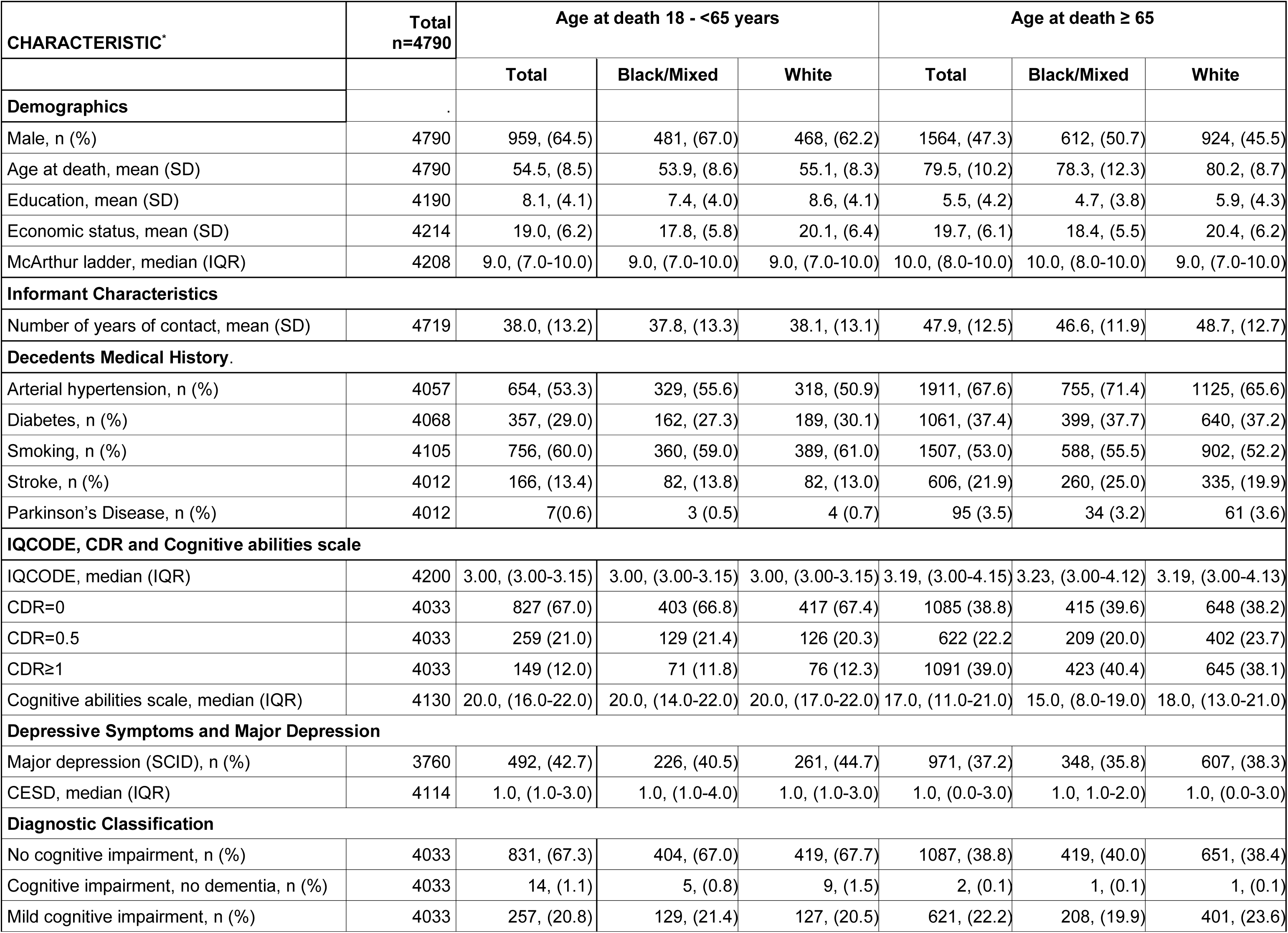
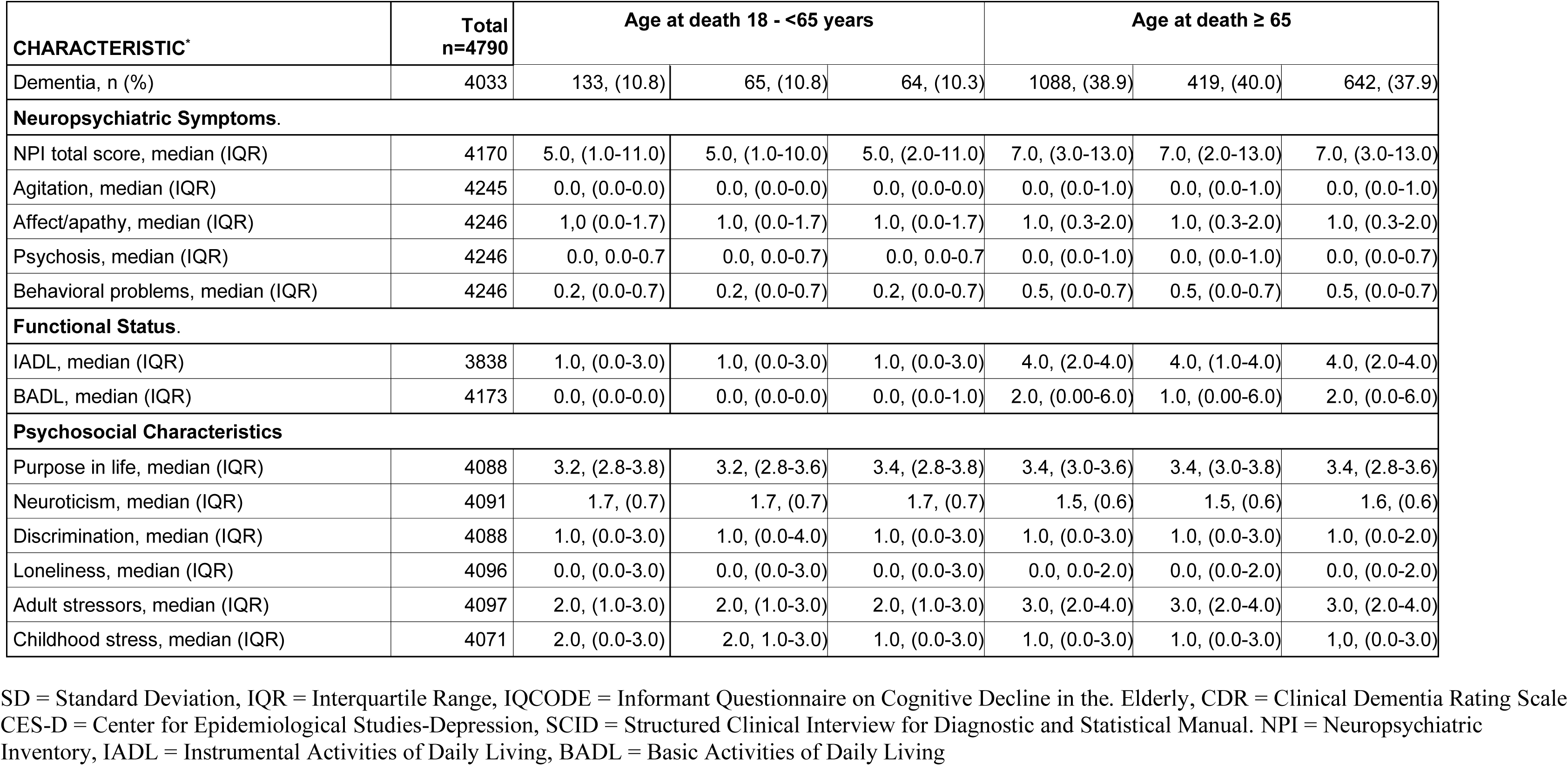
Demographics and clinical characteristics of Black/Mixed and Whites participants.

| CHARACTERISTIC* | Total<br>n=4790 | Age at death 18 - <65 years |  |  | Age at death ≥ 65 |  |  |
| --- | --- | --- | --- | --- | --- | --- | --- |
|  |  | Total | Black/Mixed | White | Total | Black/Mixed | White |
| Demographics |  |  |  |  |  |  |  |
| Male, n (%) | 4790 | 959, (64.5) | 481, (67.0) | 468, (62.2) | 1564, (47.3) | 612, (50.7) | 924, (45.5) |
| Age at death, mean (SD) | 4790 | 54.5, (8.5) | 53.9, (8.6) | 55.1, (8.3) | 79.5, (10.2) | 78.3, (12.3) | 80.2, (8.7) |
| Education, mean (SD) | 4190 | 8.1, (4.1) | 7.4, (4.0) | 8.6, (4.1) | 5.5, (4.2) | 4.7, (3.8) | 5.9, (4.3) |
| Economic status, mean (SD) | 4214 | 19.0, (6.2) | 17.8, (5.8) | 20.1, (6.4) | 19.7, (6.1) | 18.4, (5.5) | 20.4, (6.2) |
| McArthur ladder, median (IQR) | 4208 | 9.0, (7.0-10.0) | 9.0, (7.0-10.0) | 9.0, (7.0-10.0) | 10.0, (8.0-10.0) | 10.0, (8.0-10.0) | 9.0, (7.0-10.0) |
| Informant Characteristics |  |  |  |  |  |  |  |
| Number of years of contact, mean (SD) | 4719 | 38.0, (13.2) | 37.8, (13.3) | 38.1, (13.1) | 47.9, (12.5) | 46.6, (11.9) | 48.7, (12.7) |
| Decedents Medical History. |  |  |  |  |  |  |  |
| Arterial hypertension, n (%) | 4057 | 654, (53.3) | 329, (55.6) | 318, (50.9) | 1911, (67.6) | 755, (71.4) | 1125, (65.6) |
| Diabetes, n (%) | 4068 | 357, (29.0) | 162, (27.3) | 189, (30.1) | 1061, (37.4) | 399, (37.7) | 640, (37.2) |
| Smoking, n (%) | 4105 | 756, (60.0) | 360, (59.0) | 389, (61.0) | 1507, (53.0) | 588, (55.5) | 902, (52.2) |
| Stroke, n (%) | 4012 | 166, (13.4) | 82, (13.8) | 82, (13.0) | 606, (21.9) | 260, (25.0) | 335, (19.9) |
| Parkinson’s Disease, n (%) | 4012 | 7(0.6) | 3 (0.5) | 4 (0.7) | 95 (3.5) | 34 (3.2) | 61 (3.6) |
| IQCODE, CDR and Cognitive abilities scale |  |  |  |  |  |  |  |
| IQCODE, median (IQR) | 4200 | 3.00, (3.00-3.15) | 3.00, (3.00-3.15) | 3.00, (3.00-3.15) | 3.19, (3.00-4.15) | 3.23, (3.00-4.12) | 3.19, (3.00-4.13) |
| CDR=0 | 4033 | 827 (67.0) | 403 (66.8) | 417 (67.4) | 1085 (38.8) | 415 (39.6) | 648 (38.2) |
| CDR=0.5 | 4033 | 259 (21.0) | 129 (21.4) | 126 (20.3) | 622 (22.2) | 209 (20.0) | 402 (23.7) |
| CDR≥1 | 4033 | 149 (12.0) | 71 (11.8) | 76 (12.3) | 1091 (39.0) | 423 (40.4) | 645 (38.1) |
| Cognitive abilities scale, median (IQR) | 4130 | 20.0, (16.0-22.0) | 20.0, (14.0-22.0) | 20.0, (17.0-22.0) | 17.0, (11.0-21.0) | 15.0, (8.0-19.0) | 18.0, (13.0-21.0) |
| Depressive Symptoms and Major Depression |  |  |  |  |  |  |  |
| Major depression (SCID), n (%) | 3760 | 492, (42.7) | 226, (40.5) | 261, (44.7) | 971, (37.2) | 348, (35.8) | 607, (38.3) |
| CESD, median (IQR) | 4114 | 1.0, (1.0-3.0) | 1.0, (1.0-4.0) | 1.0, (1.0-3.0) | 1.0, (0.0-3.0) | 1.0, 1.0-2.0) | 1.0, (0.0-3.0) |
| Diagnostic Classification |  |  |  |  |  |  |  |
| No cognitive impairment, n (%) | 4033 | 831, (67.3) | 404, (67.0) | 419, (67.7) | 1087, (38.8) | 419, (40.0) | 651, (38.4) |
| Cognitive impairment, no dementia, n (%) | 4033 | 14, (1.1) | 5, (0.8) | 9, (1.5) | 2, (0.1) | 1, (0.1) | 1, (0.1) |
| Mild cognitive impairment, n (%) | 4033 | 257, (20.8) | 129, (21.4) | 127, (20.5) | 621, (22.2) | 208, (19.9) | 401, (23.6) |
| Dementia, n (%) | 4033 | 133, (10.8) | 65, (10.8) | 64, (10.3) | 1088, (38.9) | 419, (40.0) | 642, (37.9) |
| **Neuropsychiatric Symptoms.** |  |  |  |  |  |  |  |
| NPI total score, median (IQR) | 4170 | 5.0, (1.0-11.0) | 5.0, (1.0-10.0) | 5.0, (2.0-11.0) | 7.0, (3.0-13.0) | 7.0, (2.0-13.0) | 7.0, (3.0-13.0) |
| Agitation, median (IQR) | 4245 | 0.0, (0.0-0.0) | 0.0, (0.0-0.0) | 0.0, (0.0-0.0) | 0.0, (0.0-1.0) | 0.0, (0.0-1.0) | 0.0, (0.0-1.0) |
| Affect/apathy, median (IQR) | 4246 | 1.0 (0.0-1.7) | 1.0, (0.0-1.7) | 1.0, (0.0-1.7) | 1.0, (0.3-2.0) | 1.0, (0.3-2.0) | 1.0, (0.3-2.0) |
| Psychosis, median (IQR) | 4246 | 0.0, 0.0-0.7 | 0.0, 0.0-0.7) | 0.0, 0.0-0.7 | 0.0, (0.0-1.0) | 0.0, (0.0-1.0) | 0.0, (0.0-0.7) |
| Behavioral problems, median (IQR) | 4246 | 0.2, (0.0-0.7) | 0.2, (0.0-0.7) | 0.2, (0.0-0.7) | 0.5, (0.0-0.7) | 0.5, (0.0-0.7) | 0.5, (0.0-0.7) |
| **Functional Status.** |  |  |  |  |  |  |  |
| IADL, median (IQR) | 3838 | 1.0, (0.0-3.0) | 1.0, (0.0-3.0) | 1.0, (0.0-3.0) | 4.0, (2.0-4.0) | 4.0, (1.0-4.0) | 4.0, (2.0-4.0) |
| BADL, median (IQR) | 4173 | 0.0, (0.0-0.0) | 0.0, (0.0-0.0) | 0.0, (0.0-1.0) | 2.0, (0.00-6.0) | 1.0, (0.00-6.0) | 2.0, (0.0-6.0) |
| **Psychosocial Characteristics** |  |  |  |  |  |  |  |
| Purpose in life, median (IQR) | 4088 | 3.2, (2.8-3.8) | 3.2, (2.8-3.6) | 3.4, (2.8-3.8) | 3.4, (3.0-3.6) | 3.4, (3.0-3.8) | 3.4, (2.8-3.6) |
| Neuroticism, median (IQR) | 4091 | 1.7, (0.7) | 1.7, (0.7) | 1.7, (0.7) | 1.5, (0.6) | 1.5, (0.6) | 1.6, (0.6) |
| Discrimination, median (IQR) | 4088 | 1.0, (0.0-3.0) | 1.0, (0.0-4.0) | 1.0, (0.0-3.0) | 1.0, (0.0-3.0) | 1.0, (0.0-3.0) | 1.0, (0.0-2.0) |
| Loneliness, median (IQR) | 4096 | 0.0, (0.0-3.0) | 0.0, (0.0-3.0) | 0.0, (0.0-3.0) | 0.0, 0.0-2.0) | 0.0, (0.0-2.0) | 0.0, (0.0-2.0) |
| Adult stressors, median (IQR) | 4097 | 2.0, (1.0-3.0) | 2.0, (1.0-3.0) | 2.0, (1.0-3.0) | 3.0, (2.0-4.0) | 3.0, (2.0-4.0) | 3.0, (2.0-4.0) |
| Childhood stress, median (IQR) | 4071 | 2.0, (0.0-3.0) | 2.0, 1.0-3.0) | 1.0, (0.0-3.0) | 1.0, (0.0-3.0) | 1.0, (0.0-3.0) | 1.0, (0.0-3.0) |

SD = Standard Deviation, IQR = Interquartile Range, IQCODE = Informant Questionnaire on Cognitive Decline in the Elderly, CDR = Clinical Dementia Rating Scale CES-D = Center for Epidemiological Studies-Depression, SCID = Structured Clinical Interview for Diagnostic and Statistical Manual. NPI = Neuropsychiatric Inventory, IADL = Instrumental Activities of Daily Living, BADL = Basic Activities of Daily Living

### Demographics and socioeconomic status

Of the 4,790 decedents enrolled, nearly 70% were 65+ with a mean age of nearly 80 years. Nearly half were male, more than a third were Black, about 60% were White, and 2.0% other races, primarily Asians. Their mean education was 5.5 years. Age at death was nearly 2 years lower and educational attainment 20% lower in the Black decedents. Sex was equally distributed with slightly more males among the Black decedents.

Just over 30% decedents were age <65, the mean age was nearly 55 years, two thirds were male, about half were Black with only about 1% being primarily Asian. Their mean education was 8 years. Age at death was approximately 1 year lower and educational attainment nearly 15% lower in the Black decedents. Figure 6 shows the number of decedents by sex and race per decade.

**Figure 6.**
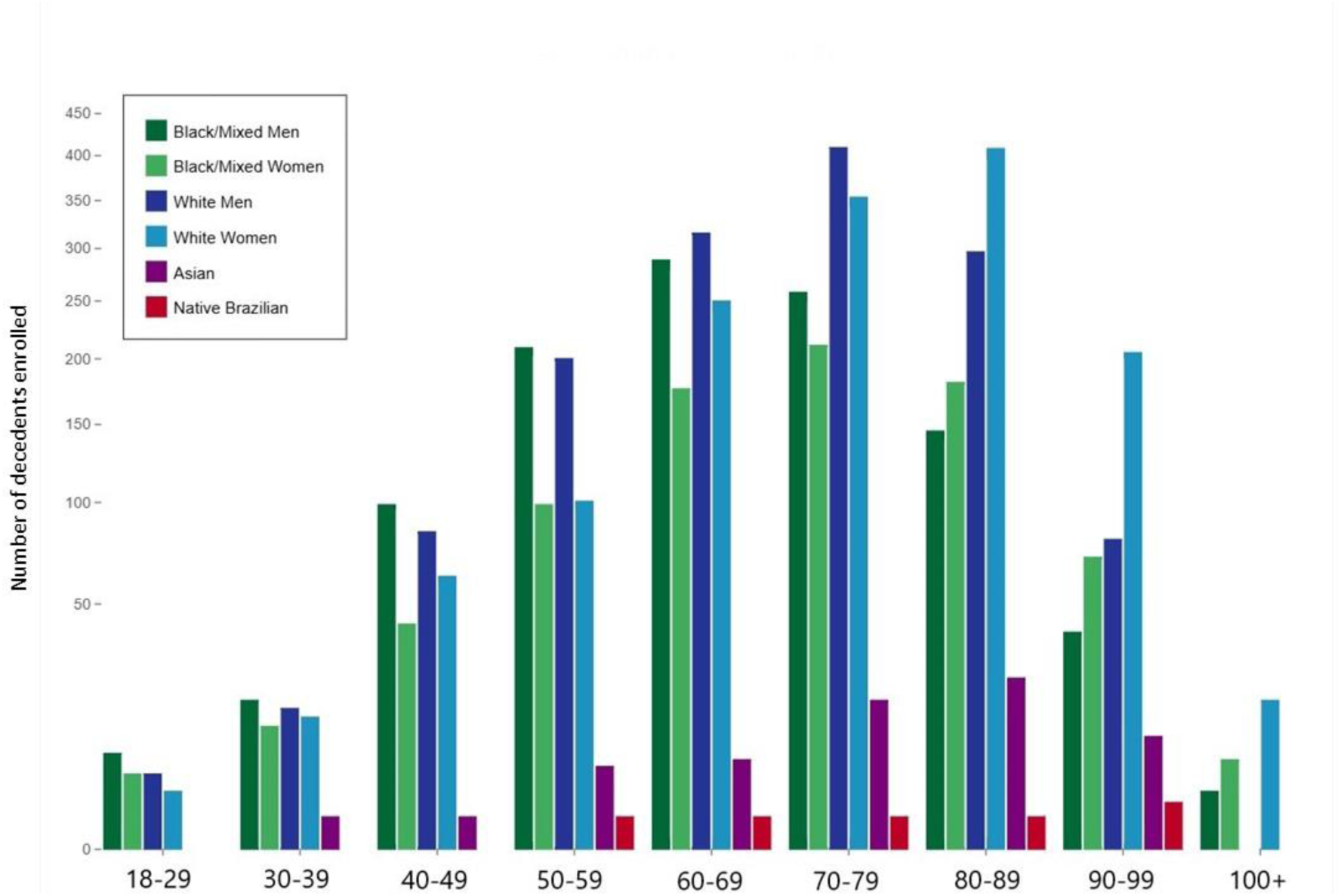
Sex and race distribution of PARDoS participants by decade from July 2021 to February 2025.

### Informant Interview

An interview was obtained from nearly 90% of informants including just over 90% for those 65+, with 116 pending, while 200 (6.1%) refused. Among those <65 nearly 90% consented to participate in the interview, with 88 pending, and only 103 (6.9%) refused (Figure 2).

### Informant characteristics

Family members accounted for 99.7% of the informants. Among those 65+, adult children were 74.4% of the informants, followed by adult grandchildren (6.5%), spouse (5.5%) and siblings (5.2%). Informants knew the decedents for an average of 44.9 years (SD: 13.5 years), nearly 50 years for the 65+ group. Informants reported having daily contact with 63.3% of the decedents, weekly in 25.8%, monthly in 8.4% and yearly in 2.4%. These frequencies were similar by race.

Among those <65, 40.4% of the informants were adult children, followed by siblings (32.8%), spouse (15.7%) and parents (2.8%). Frequencies of contact was similar in the <65 group as those in the 65+ group and were similar by race

### Socioeconomic status

In the 65+ group, the C strata was the most common in both Black and White decedents. However, the most privileged A and B strata, was about a third of the White a fifth of the Black decedents and the most vulnerable D and E strata, were more than 15% of the Black and just over 10% of the White decedents. Interestingly, these differences were not found in the social status measured by the McArthur ladder.

Similarly, among the <65 group, C strata was also the most common for both Black and White decedents. A and B strata was in a fifth of the Black while more than a third in the White decedents, and D and E strata was in more than a fifth of Black but just over 10% of White decedents.

### Decedents Medical History

Diabetes was present in approximately a third, arterial hypertension in approximately two thirds, and smoking in more than half of the 65+ group. Stroke was found in a nearly a quarter and Parkinson’s disease in 3.5%. Older Black had 10% more hypertension and 25% more stroke than White decedents.

Among the <65 group, diabetes was found in a quarter, hypertension in half and smoking in nearly two thirds of the decedents. Stroke was present in just over 10% and Parkinson’s disease in 0.6% of decedents.

### IQCODE and CDR

The median IQCODE was 3.2 (IQR: 3.0-4.2) for 65+ group. An ICQODE > 3.3 was found in 45.6% of the 65+ group. Cognitive impairment, i.e., CDR>0 was present in just over 60% of the 65+ participants, including about 40% with CDR>0.5. The IQCODE is used as a measure of cognitive decline.

The median IQCODE was 3.0 (IQR: 3.0-3.2) for the <65 group. An ICQODE > 3.3 was found in 18% of the 18-<65 participants, and a third had CDR>0 including 12% with CDR>0.5.

### Cognitive abilities and Literacy

In the 65+ group, White decedents had acquired approximately 20% more cognitive abilities than Black decedents. Verbal literacy and numeracy were 20% greater in the White compared to the Black decedents. The frequencies for music literacy were similar for Black and White decedents.

The median number of acquired cognitive abilities was similar for Black and White decedents in the <65 group. Verbal literacy was 10% greater in White while numeracy and music literacy were similar among Black and White decedents.

### Depressive Symptoms and Major depression

Major depression in the past was present in more than a third of the 65+ group and was approximately 10% higher in the White compared to the Black decedents. Median CESD was similar between both Black and White groups.

Among the <65 decedents, major depression was found in more than 40%, and again 10% more frequent in White compared to Black decedents. CESD was similar in both Black and White decedents.

### Diagnostic Classification

In the 65+ group, MCI was present in approximately 20% participants, with just over 80% being amnestic. Dementia was found in a third of the 65+ participants, with 96% classified as Alzheimer’s dementia with or without other contributing conditions.

Vascular cognitive impairment (VCI) was present in 4.2%, whereas Parkinson’s/ Parkinsonism related cognitive impairment was present in 1.8% of participants. The diagnosis of CIND was found in only 2 participants, both of whom had mental retardation. The frequency of MCI was nearly 4% higher in White decedents while dementia was 2% higher in the Black decedents unadjusted for age and education. The distribution of the clinical diagnosis among races is shown in Figure 7.

**Figure 7.**
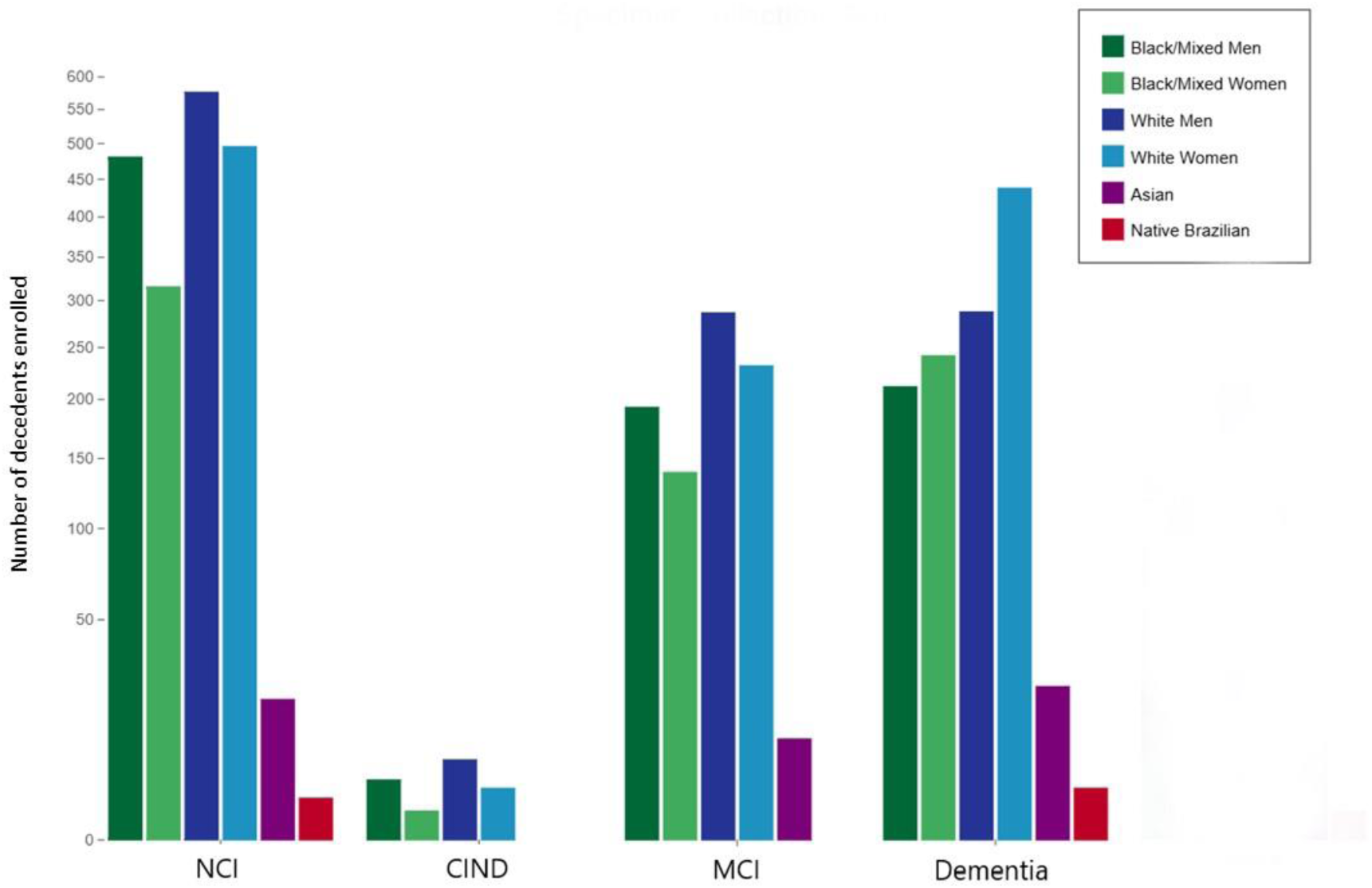
Distribution of the clinical diagnosis among races. NCI = No cognitive impairment, CIND = Cognitive impairment with no dementia, MCI = Mild cognitive impairment

Other contributors to dementia included alcohol abuse in nearly a quarter of 65+ decedents, seizures in 8.0%, dialytic chronic kidney disease in 5%, bipolar disease in less than 5%, schizophrenia in almost 3%, drug abuse in almost 2% and brain tumors in 0.5%. There were 6 decedents with HIV infection and 1 with Down Syndrome. None of the cases had autism, microcephaly, anoxic encephalopathy or multiple sclerosis.

In <65 group, MCI was found in 20%, with more than 80% being amnestic. Dementia was present in 10%, with more than 85% being Alzheimer’s dementia. VCI was present in 7.9% and Parkinson’s/Parkinsonism related cognitive impairment was present in 1.8% related dementia. CIND was found in 14 participants, with mental retardation in 5, hydrocephalus in 3, anoxic encephalopathy in 2 and other conditions in 4 participants.

In the <65 group, alcohol abuse was reported in about 40%, drug abuse and seizures in nearly 15%, bipolar disease and dialytic chronic kidney disease in 5% each, schizophrenia in 4%, HIV infection in 1.0%. Brain tumors were reported in 8 decedents, Down Syndrome and multiple sclerosis in 2 decedents each.

### Neuropsychiatric symptoms

The median NPI was 7.0 for the 65+ group. The NPI global scores and the scores of each of the four clusters of symptoms were similar among the Black and White decedents. Affect/apathy was the most frequently affected domain in both Black and White decedents, followed by the behavioral problem domain.

Among the <65 group, the median NPI was 5.0. Global scores and domains were similar among Black and White decedents and affect/apathy was also the most commonly affected domain.

### Functional status

Median IADL and BADL were 4.0 and 2.0 for 65+ group, respectively. While IADL scores were similar among races, BADL median scores were 50% lower in Black compared to White decedents.

For the <65, median IADL and BADL were 1.0 and 0, respectively. Both IADL and BADL were similar among Black and White decedents.

### Other scales

Among the 65+ participants, nearly half of the White and more than half of the Black decedents reported suffering from discrimination. Neuroticism was slightly higher in White compared to Black decedents. Scores for purpose in life, loneliness, childhood and adult stressors were similar among both races.

For the <65 participants, discrimination was 10% more frequent in Black than White decedents. Median childhood stress scores were greater in Black compared to White decedents. Similar scores for purpose in life, loneliness, neuroticism and adult stressors were found in both Black and White decedents.

## Discussion

PARDoS is collecting brains and clinical data from an informant from a large and diverse sample of Brazilians to identify the environmental, genetic and molecular drivers of ADRD clinical and neuropathologic traits. Clinical-pathologic studies have been instrumental to our understanding of ADRD. Understanding the importance of these pathologies requires having comparable persons with and without dementia. This is best accomplished by including autopsy as part of community-based studies of aging and dementia. A number of such studies are ongoing with variable autopsy rates.[19–34] PARDoS complements the other ongoing studies in terms of age range, race, SES and the availability of other organs from a large number of Brazilians.

Most autopsies from community-based clinical-pathologic studies are almost exclusively conducted in non-Latinx White decedents. There is a study, also based in Brazil, primarily of older persons, which recruited Blacks and Whites decedents.[33,34] PARDoS complements the portfolio of clinical-pathologic studies in several ways.

Through April 10, 2025 we collected 5034 brains with about 6 brains being collected per day, we anticipate at least 6500 autopsies by the end of 2025. More than 40% are Black decedents given the enrichment for African ancestry. PARDoS is already generating APOE and whole genome sequencing for all participants. and subsets with other omics with a goal of generating a large matrix of multi-omic data similar to ROSMAP in a larger and more diverse sample.[32,82] Further, a third of PARDoS brains are person ages 18-64. Finally, samples from other organs were collected from a subset of participants.

Epidemiological studies report that African Americans and Latinxs are more likely to have dementia than non-Latinx Whites.[83–86] Previous studies showed race differences in the prevalence of environmental and genomic risk factors for dementia, and differences in the association of these risk factors with dementia among different races.[87–96] For example, apolipoprotein ε4 allele (APOE ε4), the most important genetic risk factor for AD in non-Latinx White participants, is more common among African Americans but its association with Alzheimer’s dementia risk in African Americans is weaker.[97–99] Recent Alzheimer’s dementia genome-wide association study (GWAS) of African-American and Latinx populations found unique variants and failed to replicate some variants found in non-Latinx white participants.[82,100–104] PARDoS is well-positioned to extend this work to understand the influence of African ancestry on the relation of genomic and other molecular drivers of ADRD neuropathologic traits.

Most autopsies from community-based clinical-pathologic studies are almost exclusively older individuals with very few autopsies under the age of 70 years. PARDoS complements the ongoing work with about a third of autopsies being persons aged <65. Over the last several years PET, CSF, and now blood biomarkers suggest that AD begins long before clinical symptoms.[105–114] Much less is known about other neurodegenerative diseases which are currently beyond the resolution of *in vivo* biomarkers. Brains of younger participants are essential to study the earliest molecular changes that occur before and with the onset of AD and other neuropathologies. Thus, PARDoS can inform on strategies for the primary prevention of ADRD neuropathologic traits.

Most autopsies from community-based clinical-pathologic studies are almost exclusively done in highly educated individuals with a high socioeconomic status (SES). PARDoS complements the ongoing work with an average of 6 years of education and the full range of education from illiterate to post-graduate and the full range of SES. Epidemiological studies report that lower education and lower SES are associated with a higher risk of dementia.[115–119] The illiterate individuals are most vulnerable and even a few years of education may have an impact in reducing the risk of dementia.[120,121] We and other have demonstrated that education and SES may modify the association of AD and other pathologies with dementia.[122–125] PARDoS offers a platform to more fully understand the relation of SES to ADRD neuropathologic traits.

Most autopsies from community-based clinical-pathologic studies are limited to brain. PARDoS complements the ongoing work; since a full-body autopsy is done and other organs can be obtained for research. PARDoS is approved to collect a variety of tissues other than the brain and can create a unique resource to investigate brain-body interactions associated with ADRD. There is consistent evidence demonstrating that AD and other common neuropathologies are not restricted to the brain. β-amyloid is produced in peripheral tissues or cells, such as platelets, skin fibroblasts, skeletal muscles, and cardiovascular smooth muscle cells, and is secreted into the blood circulation [126–128] Lewy bodies are also found in peripheral nerves and plexuses, submandibular gland, muscles and fat, as well as the brain.[129–131] Thus access to these tissues could provide yet another valuable asset to PARDoS.

The major limitation of PARDoS is that there is no prospective data collection, and the clinical data are mostly limited to an informant interview. For cases obtained at IAMSPE, we can link to their electronic medical records which have their own limitations. However, the full-body autopsy offers us the opportunity to capture a range of lifetime exposure data, the exposome, in ways that complement and, in some cases, improve on what can be done in living persons. For example, teeth and bone capture early-life and cumulative metal exposure.[132] Lung and bronchus capture lifelong exposure to 2.5 particulate matter.[133] Abdominal and pericardial fat and liver are depots for persistent organic pollutants.[134]. Other organs harbor microplastics. Given the large number of brains collected daily, the protocols for neuropathologic data collection were streamlined in comparison with other tertiary care dementia centers across the globe. A number of steps were introduced to make the high throughput data collection feasible.

## Data Availability

PARDoS data can be requested online at www.radc.rush.edu

## Acknowledgement

The authors thank the thousands of Brazilian representatives who participated in this study and PARDoS staff in Brazil and USA. We thank the Rush Alzheimer’s Disease Center, the Núcleo de Estudos, Pesquisa e Assessoria à Saúde (NEPAS), the Autopsy Services (SVO) at Santo André and Guarulhos, and the Instituto de Assistência Médica ao Servidor Público Estadual (IAMSPE).

PARDoS is supported by NIA grants R01AG54058 and R01AG075927.

The data repository is at the RADC and requires a Data Use Agreement (DUA) with RUMC which includes language to make it compliant with Brazilian law. The PARDoS biorepository is in IAMSPE. Material can only be sent abroad or used with money from a foreign country with approvals from a Brazilian local ethical committee (CEP) and the national ethical committee (CONEP). Such approvals can only be granted to a Brazilian affiliated with an institution in Brazil. Thus, access to PARDoS resources can only be done in collaboration with a Brazilian investigator and institution. A Material Transfer

Agreement (MTA) will be required with IAMSPE. The DUA and MTA are tailored to Brazilian laws and regulations and cannot be negotiated. Persons interested in accessing PARDoS resources will need to work with their institution to ensure the DUA and MTA is accepted as written.

PARDoS resources can be requested at www.radc.rush.edu.

The investigators have no relevant conflicts of interest. The funder had no role in the generation of this manuscript.

## Supplemental Text

### Data Management

The process starts with the generation of a unique and random 10-digit alpha-numeric identifier with a check digit for each subject. The intake workflow contains the intake of demographic data from the recruiting sites, the verification of a legal informant, and obtaining study consent.

Nurse interviewers enter the demographic data in the intake form. As representatives of more than one decedent often arrive about the same time, the interviewers first obtain the proxy-reported race, education and place of birth as these variables are used to identify high priority cases.

Upon obtaining consent, information becomes available to the nurse interviewers and the autopsy staff. Completion of the intake form initiates the collection of data from the informant by the nurse interviewer, specimen collection and processing by the autopsy technician and specimen storage by the laboratory staff, and subsequent neuropathologic evaluation.

### Aging and Dementia Multiracial Genetic Cohort (ADMIX)

From January 26th, 2021 to March 16th, 2023, ADMIX recruited 1046 decedents age 18 years or older. Their mean age of death was 75.5 years (SD: 12.7 years, range 24 to 103), 44.0% were male, 31.9% were Black/Mixed, 65.5% were Whites and mean education was 7.9 years (SD: 5.1 years). ADMIX was discontinued to devote the staff time to obtaining interviews for those on whom we obtained the brain.

### Study of Ancestry Neurodegenerative Diseases and Stroke (SANDS)

The initial grant named African Ancestry and the Genomic Architecture of AD and Other Common Neurodegenerative Disease Neuropathologies (R01AG054058) was awarded to Dr. Bennett and Rush University Medical Center (RUMC) on 15 September 2016 and conducted activities at the Faculty of Medicine University of Sao Paulo (FMUSP) with Dr. Farfel as the subcontract principal investigator from November 2016 through July 2018 under the name Study of Ancestry Neurodegenerative Diseases and Stroke (SANDS). On September 5th, 2019, FMUSP notified RUMC that they would no longer support the project. Subsequently, approvals were received from CEP of IAMSPE on May 12th, 2020 and from CONEP on June 13th, 2020 to move SANDS from FMUSP to IAMSPE. SANDS remains active and approved in CONEP, but data and biospecimen collection had stopped in July 2018and was never resumed.

### Ethical Approvals in Brazil

PARDoS was submitted to CEP and CONEP as a new study, using a specific consent and was approved by CEP on May 12, 2020 and by CONEP in September 17, 2020, more than two years after we discontinued SANDS. We initially planned to merge SANDS with PARDoS and published seven papers under the name PARDoS with data from both FMUSP and IAMSPE. [54,60,62,69,135–138] However, we subsequently elected to separate SANDS from PARDoS. The decision stemmed in large part from the fact that the procedures at IAMSPE, Santo Andre and Guarulhos SVOs allowed for much faster access to the brains, improved processing protocols, and that we could obtain other organs for substudies all of which make the quality of the brain and other biospecimens collected at IAMSPE distinct from SANDS. All work with SANDS data ceased on September 19, 2022. The paper with a publication date of 2024 was submitted in its final form on April 30, 2021.[136]

